# Neural correlates of adverse childhood experiences, depression and chronic pain in the reward system

**DOI:** 10.1101/2025.07.10.25331260

**Authors:** Georgia Antoniou, Blair H. Smith, Liana Romaniuk, Heather C. Whalley, Andrew M. McIntosh, Gordon D. Waiter, Tim G. Hales, J. Douglas Steele, Lesley A. Colvin

## Abstract

Adverse childhood experiences (ACEs) are common, altering behaviour and stress reactivity, with persistent structural and functional brain changes. ACEs are associated with a greater risk of physical and mental health morbidities, including depression and chronic pain (CP), which often co-exist. While changes in the reward system have been implicated in each condition, it remains unclear whether there are common neural mechanisms. Using brain scans from a large community sample, we examined how ACEs, depression, and CP affected the reward system. Individuals reporting both ACEs and CP exhibited reduced volume in the nucleus accumbens, a brain region associated with motivation and pleasure, particularly among women. Blunted reward-related activity in the basal ganglia was linked to higher depression scores, CP severity, and experiences of childhood sexual abuse. Importantly, these functional changes remained significant even after accounting for structural brain differences. Interestingly, brain structural-functional associations were weak, suggesting distinct underlying mechanisms. Structural brain alterations likely represent cumulative, enduring consequences of ACEs and CP, whereas altered reward activity appears more closely tied to current symptom severity. These findings reveal separable but converging neurobiological signatures of early adversity, pain, and depression within the brain’s reward system.

## Introduction

Adverse childhood experiences (ACEs) are events or situations likely to cause harm or distress that a child either experiences themself or encounters within their environment, which undermine their sense of safety, with approximately half of all children having experienced at least one ACE^1^. Examples include physical abuse/neglect, emotional abuse/neglect, sexual abuse, discrimination, poor health, substance misuse in the home, war, loss of a parent, and bullying. There are potential long-term effects of ACEs on stress responses, with a greater risk of physical and mental health morbidity and mortality in adulthood^2,3^.

ACEs are considered a risk factor for developing depressive illness later in life and are associated with an elevated risk of multimorbidity, including both mental and physical health problems^4^. Depression is highly prevalent in individuals with chronic pain (CP), with co-morbidity rates reaching as high as 66.3%^5^. CP itself is a leading global cause of years lived with disability, with its prevalence ranging from 35% to 53%^6–10^. This raises an important question: is the co-existence of depression, CP and ACEs due to their high prevalence, or is there a shared underlying mechanism driving these associations? Although many studies report associations between these occurrences, the exact pathways linking them remain unclear.

Whilst there is evidence of persisting central changes in adults with depression who have a history of ACEs, there is a significant gap in the existing literature, investigating the neural substrates for CP and ACEs^11^. There are no studies investigating the central changes associated with co-morbid depression and CP with a history of ACEs^11^. Anhedonia is a core feature of depressive illness, and many studies on depression report blunted brain reward event-related activations and brain structure changes^12^, with reduced striatal reward activation associated with more severe depression^12–23^. Independently of studies on depression, blunted reward-linked striatal activity is also associated with pain and CP conditions^24^. These alterations in reward processing, may have shared mechanisms in CP and depression, although the association with neural changes resulting from ACEs are poorly understood^24^. Regarding ACEs, there is evidence that early-age maltreatment or neglect is more likely to lead to a reduction in striatal responses to rewarding stimuli later in life^25^.

ACEs can affect the brain at multiple levels, including changes at epigenetic, cellular, systems and network levels^26–28^. Whilst there is evidence that CP, depression and ACEs are linked, a detailed understanding of common mechanisms and moderators remains unclear^11^. Alterations in the dopaminergic system within the striatum in CP conditions^29^, coupled with the role of the nucleus accumbens (NAcc) in mood disorders and reward processing, point to a potential common pathway^30^. Nevertheless, the available studies examining CP and ACEs independently have revealed the involvement of several brain regions, including the striatum, insula, amygdala, and hippocampus.

Given the complex interplay between ACEs, depressive symptoms, and CP in adulthood, this study aimed to examine their shared and unique neural correlates, with a particular focus on reward-related brain systems. We hypothesised that these clinical factors would be associated with both structural and functional alterations in reward-related brain regions. In particular, we expected that structural and functional changes would contribute independently to these clinical factors, consistent with the idea that ACEs, CP, and depression exert their influence on neural systems through partially dissociable, yet interacting, pathways.

## Results

A total of 1,080 participants were initially considered for inclusion in the analyses. After quality control, 32 participants were excluded due to inadequate segmentation quality and an additional 201 were excluded due to missing questionnaire data, resulting in a final sample of 847 participants for the structural MRI analyses (Group 1; **Table 1**). This group included 525 females (61.98%), with a mean age of 59.03 years (SD = 10.33; range 26–84). The sample was predominantly white (840/847), with two participants identifying as mixed race and five with missing ethnicity data (**Table 1**).

**Table 1:** Clinical and Demographic details of the population for the structural MRI analysis. Includes associations of age, sex, depression, and adverse childhood experiences with ‘chronic pain’. Age was assessed during face-to-face entry of the STRADL study. Quick Inventory of Depressive Symptomatology; QIDS, chronic pain grade; CPG, adverse childhood experiences; ACEs.

|  |  | Subgroup category | Proportion of participants with CP | median | se | min | max | mean | sd |
| --- | --- | --- | --- | --- | --- | --- | --- | --- | --- |
| Age (years) |  | - | - | 61 | 0.35 | 26 | 84 | 59.03 | 10.33 |
|  |  | 25-34 | 15/32 | - | - | - | - | - | - |
|  |  | 35-44 | 15/57 | - | - | - | - | - | - |
|  |  | 45-54 | 40/118 | - | - | - | - | - | - |
|  |  | 55-64 | 198/424 | - | - | - | - | - | - |
|  |  | 65-74 | 87/189 | - | - | - | - | - | - |
|  |  | >75 | 13/27 | - | - | - | - | - | - |
| Sex* |  | F | 245/525 | - | - | - | - | - | - |
|  |  | M | 123/322 | - | - | - | - | - | - |
| Chronic Pain | CPG | 0 (No pain) | 368/847 | 1 | 0.002 | 1 | 4 | - | - |
|  |  | I (low-disability-low intensity) | 220/368 | - | - | - | - | - | - |
|  |  | II (low-disability-high intensity) | 95/368 | - | - | - | - | - | - |
|  |  | III (high disability-moderately limiting) | 28/368 | - | - | - | - | - | - |
|  |  | IV (high disability-severely limiting) | 25/368 | - | - | - | - | - | - |
| Depression | Severity of Depression | - | 47/64 | 0 | 0.001 | 0 | 4 | - | - |
|  |  | 0 | 234/613 | - | - | - | - | - | - |
|  |  | 1 | 87/170 | - | - | - | - | - | - |
|  |  | 2 | 23/39 | - | - | - | - | - | - |
|  |  | 3 | 20/21 | - | - | - | - | - | - |
|  |  | 4 | 4/4 | - | - | - | - | - | - |
|  | QIDS | - | - | 3 | 0.1 | 0 | 22 | 4.64 | 3.71 |
| Adverse Childhood Experiences, (Childhood Trauma Questionnaire sub-scores) | Physical Abuse | - | 65/112 | 5 | 0.003 | 5 | 25 | - | - |
|  | Sexual Abuse | - | 43/75 | 5 | 0.004 | 5 | 25 | - | - |
|  | Emotional Abuse | - | 72/119 | 5 | 0.004 | 5 | 25 | - | - |
|  | Physical Neglect | - | 102/182 | 5 | 0.003 | 5 | 21 | - | - |
|  | Emotional Neglect | - | 61/90 | 7 | 0.005 | 5 | 25 | - | - |
|  | Denial | - | - | 1 | 0.001 | 0 | 3 | - | - |
| Multiple ACEs | No ACEs | - | 210/543 | - | - | - | - | - | - |
|  | One ACE | - | 71/170 | - | - | - | - | - | - |
|  | Two ACEs | - | 34/57 | - | - | - | - | - | - |
|  | Three ACEs | - | 22/32 | - | - | - | - | - | - |
|  | Four ACEs | - | 17/27 | - | - | - | - | - | - |
|  | Five ACEs | - | 14/18 | - | - | - | - | - | - |
| Total Intracranial Volume TIV (cm<sup>3</sup>) |  | - | - | 1436.5 | 0.163 | 1110 | 877 | - | - |

Among this cohort, a subgroup of 604 participants met the inclusion criteria, which included 99% coverage of the basal ganglia in the task-based fMRI data and completion of both the Quick Inventory of Depressive Symptomatology (QIDS) and the Childhood Trauma Questionnaire (CTQ) (Group 2; **Table 2**). This subgroup included 392 females (64.90%), with a mean age of 58.15 years (SD = 10.99; range 26–84), and a racial composition similar to Group 1. Among them, 252 participants reported chronic pain (CP) at baseline and had completed the Chronic Pain Grade (CPG) questionnaire.

**Table 2:**
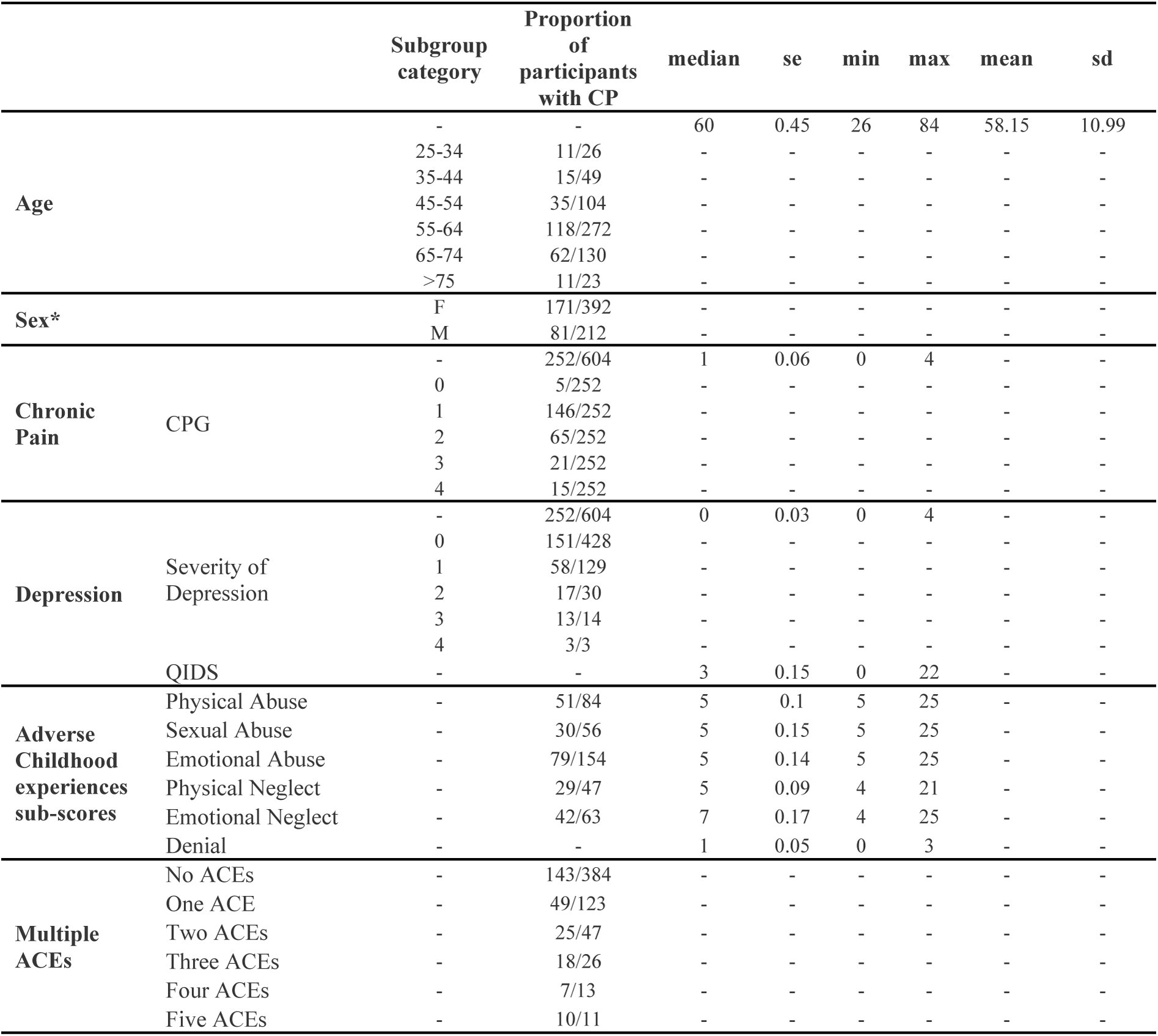
Clinical and demographic characteristics of the population for the task-based fMRI analysis. Includes associations of age, sex, depression, and adverse childhood experiences with ‘chronic pain’. Age was assessed during face-to-face entry of the STRADL study. Quick Inventory of Depressive Symptomatology; QIDS, chronic pain grade; CPG, adverse childhood experiences; ACEs.

A final subset of 552 participants had both structural and functional MRI data (Group 3; **Table 3**). This group comprised 378 females (68.47%), with a mean age of 58.07 years (SD = 11.01; range 26–84), and the racial composition remained consistent across groups. Analyses in this subgroup were used to examine associations between functional reward system activation and structural differences in corresponding brain regions.

**Table 3:**
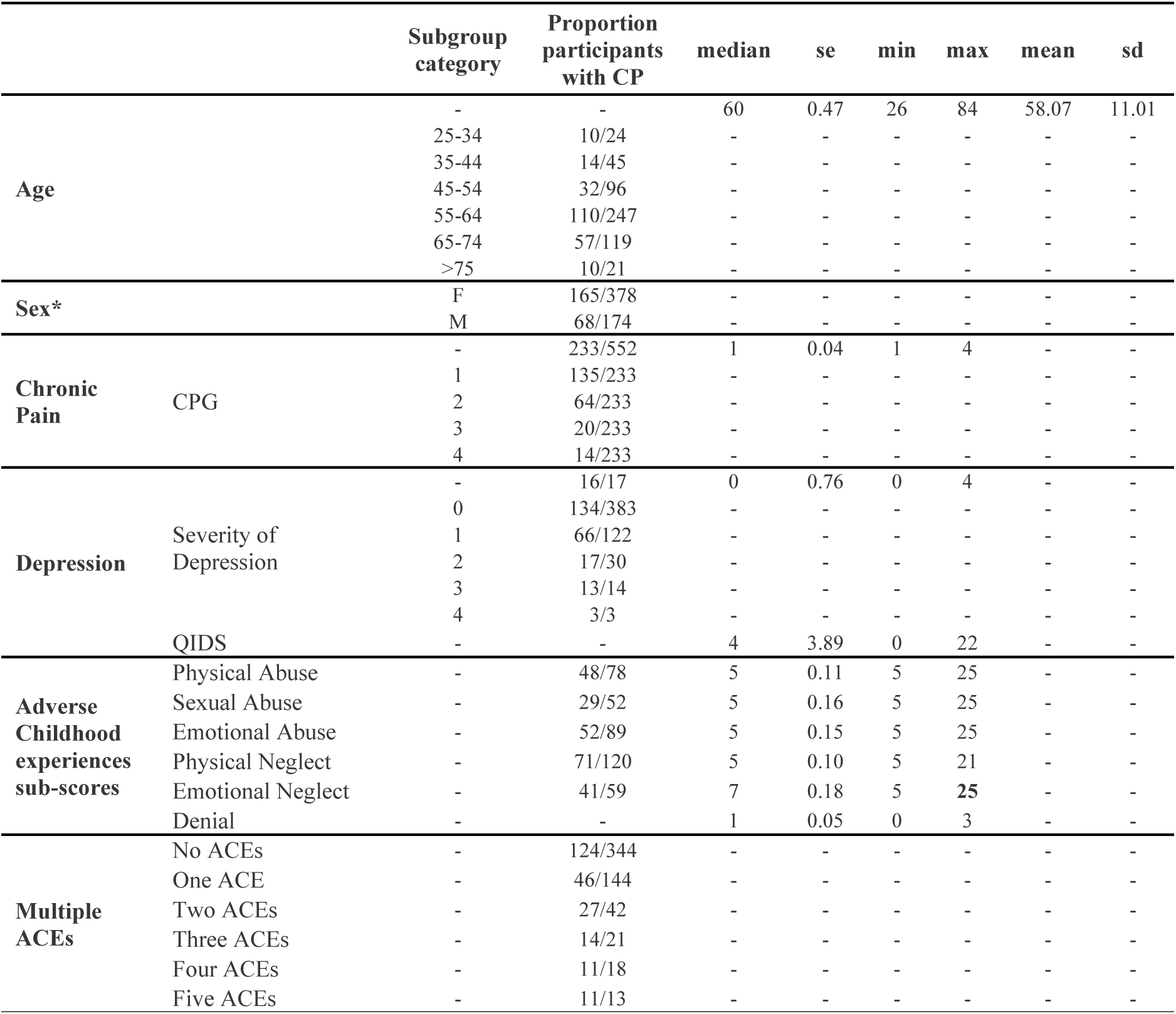
Clinical and demographic characteristics of the population used in the task-based fMRI and structural analyses. Includes associations of age, sex, depression, and adverse childhood experiences with ‘chronic pain’. Age was assessed during face-to-face entry of the STRADL study. Quick Inventory of Depressive Symptomatology; QIDS, chronic pain grade; CPG, adverse childhood experiences; ACEs.

### Predictors of Chronic Pain

A logistic regression model was used to identify predictors of CP. ACEs, age, depressive symptoms (QIDS), and anxiety symptoms (Hospital Anxiety and Depression Scale; HADS) all emerged as significant predictors (**Table 4**). Each additional ACEs was associated with a 21.4% increase in the odds of CP (OR=1.21, 95% CI 1.06-1.39, p<.01), while each additional year of age increased the odds by 2.6% (OR=1.03, 95% CI 1.01-1.04, p<.001). Higher depressive symptom scores (QIDS) and anxiety scores (HADS) also predicted increased odds of CP (QIDS: OR=1.11, 95% CI 1.05-1.17, p<.001; HADS: OR=1.06, 95% CI 1.00-1.12, p<.05). These results underscore the contribution of both psychological and demographic factors to the risk of CP, although the potential for multicollinearity among these predictors requires consideration.

**Table 4:** Logistic regression results predicting chronic pain. The coefficients represent the association of each predictor variable with the likelihood of experiencing chronic pain. Significant relationships are indicated by asterisks (*p < 0.5; **p < 0.01; ***p < 0.001). The logistic regression model for chronic pain includes variables for adverse childhood experiences (ACEs), age, sex, total intracranial volume (TIV), depression severity (QIDS), and anxiety scale (HADS)

| **Logistic Regression Results** |  |  |  |  |
| --- | --- | --- | --- | --- |
| Dependent variable: Chronic Pain |  |  |  |  |
|  | **Model 1** | **Model 2** | **Model 3** | **Model 4** |
|  | **Value (SD)** | **Value (SD)** | **Value (SD)** | **Value (SD)** |
| **ACEs** | 0.321***<br>(0.063) | 0.315***<br>(0.064) | 0.199**<br>(0.068) | 0.194**<br>(0.068) |
| **Age** | 0.018**<br>(0.007) | 0.018*<br>(0.007) | 0.024**<br>(0.007) | 0.026***<br>(0.007) |
| **Sex** | 0.333*<br>(0.147) | 0.028<br>(0.195) | -0.028<br>(0.199) | -0.049<br>(0.2) |
| **TIV** |  | -0.002*<br>(0.001) | -0.002*<br>(0.001) | -0.002*<br>(0.001) |
| **QIDS** |  |  | 0.137***<br>(0.024) | 0.101***<br>(0.029) |
| **HADS** |  |  |  | 0.059*<br>(0.028) |
| **Constant** | -1.774***<br>(0.444) | 0.838<br>(1.175) | -0.13<br>(1.217) | -0.416<br>(1.228) |
| **Observations** | 847 | 847 | 847 | 847 |
| **Log Likelihood** | -559.43 | -556.53 | -537.4 | -535.23 |
| **Akaike Inf. Crit.** | 1,126.85 | 1,123.07 | 1,086.80 | 1,084.46 |

\*p<0.05; \*\*p<0.01; \*\*\*p<0.001

### Reward System Activation in Task-based fMRI

Robust activation of the reward system was observed across all 604 participants in response to task-based reward stimuli, as determined by a cluster-extent threshold for significance. Key regions showing significant activation included the putamen, caudate, pallidum, ventral striatum, dorsolateral superior frontal gyrus, medial superior frontal gyrus, and anterior cingulate cortex (ACC). Notably, blunted activation in a subset of these key regions was identified in association with CP, greater CP severity, elevated depression severity, and exposure to subtypes of ACEs. For region-specific effects, see supplementary materials.

Multiple regression analyses within a predefined basal ganglia mask (**Figure 1**a) revealed significant associations between reward-related activation (captured via the principal eigenvariate of a basal ganglia mask) and individual differences in symptomatology. Specifically, basal ganglia reward responses were inversely associated with chronic pain severity, as measured by the CPG scale (**Figure 1**b). A significant negative association was also found between basal ganglia activation and the sexual abuse subscale of the CTQ, with no significant effects observed for other CTQ subscales (**Figure 1**c). Moreover, reduced basal ganglia reward activation was significantly associated with greater depression severity, as indexed by the QIDS (**Figure 1**d).

**Figure 1:**
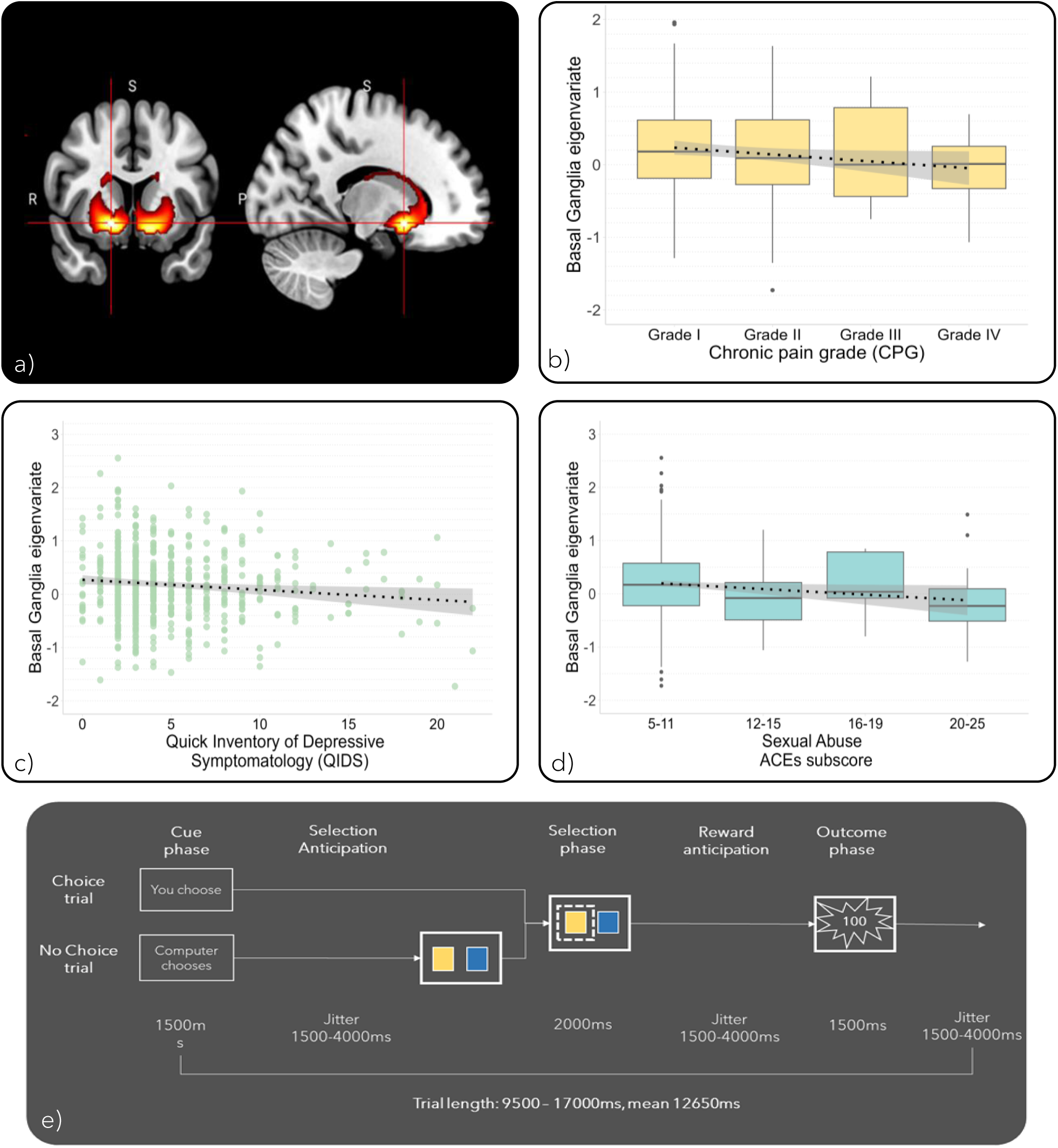
Correlations between chronic pain grade scores, sexual abuse sub-scores and depressive symptom scores, with reward signal. Higher depression severity, chronic pain severity and history of childhood sexual abuse were associated with lower basal ganglia reward response. **a)** Reward activation signal in the basal ganglia mask. **b)** Negative^)^ correlation of CPG scores with eigenvariate scores of the striatal activity mask (Spearman’s p= −0.099, P < 0.05). **c)** Negative correlation of QIDS severity scores with eigenvariate scores of the striatal activity mask (Spearman’s p= −0.082, P<0.05). **d)** Negative correlation of sexual abuse CTQ sub-scores with eigenvariate scores of the striatal activity mask (Spearman’s p= −0.011, P<0.05). **e)** Probabilistic reward learning task: The first phase of the task was a cue indicating whether the computer would choose the selection (no choice trial). If so, a participant was obliged to follow the choice. If not, the participant was able to make the choice (choice trial). The final phase was the presentation of the outcome, either ‘no reward’ (0 points) or ‘reward’ (100 points).

### Grey Matter Volume Alterations (VBM)

Voxel-based morphometry (VBM) analysis revealed grey matter alterations in reward-related brain regions, specifically the nucleus accumbens (NAcc) and Brodmann Area 25 (BA25), in individuals who reported chronic pain and different ACEs scores (**Figure 2**). Notably, this pattern did not hold for individuals with a history of sexual abuse and later-life CP, indicating a potentially distinct neural profile in this subgroup.

**Figure 2:**
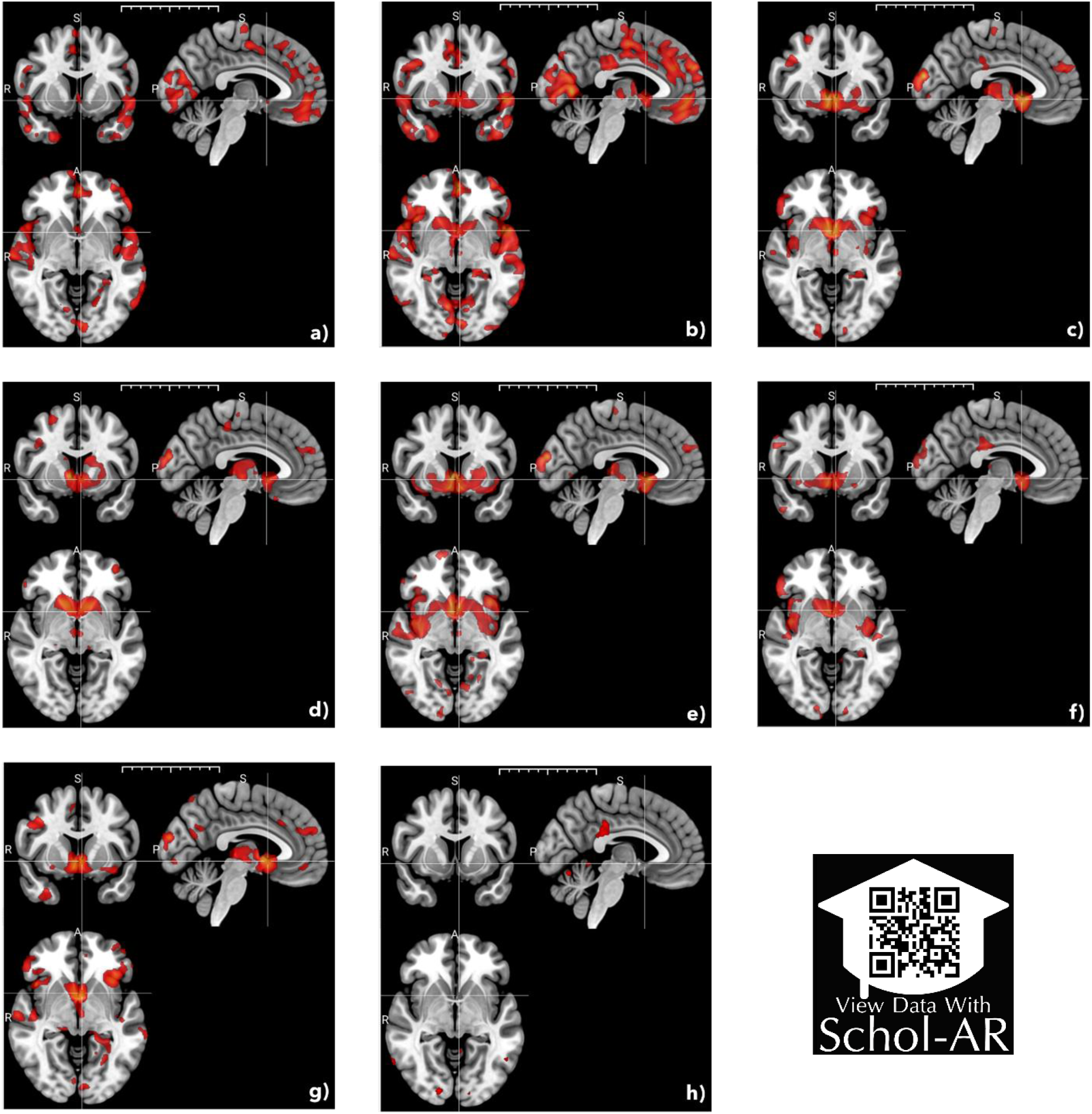
Brain images of negative association between grey matter with individuals who reported chronic pain and different Adverse childhood experience scores, Voxel Based Morphometry analysis. a) Chronic Pain vs No Chronic Pain, b) Chronic Pain severity Vs No Chronic pain, c) Chronic pain and ACEs doses Vs Healthy Controls, d) Chronic Pain and Physical abuse dual exposure Vs no dual exposure, e) Chronic Pain and Emotional abuse dual exposure Vs no dual exposure, f) Chronic Pain and Emotional neglect dual exposure Vs no dual exposure, g) Chronic Pain and Physical neglect dual exposure Vs no dual exposure, h) Chronic Pain and Sexual abuse dual exposure Vs no dual exposure.

### Structural Differences in the Nucleus Accumbens

ANCOVA models examining NAcc volume in relation to CP and ACEs exposure, adjusting for age, sex, and scan site, demonstrated a significant main effect of group (F(3,840) = 4.37, p = 0.005) (**Figure 3**) alongside strong effects of age (p < 1e-19) and sex (p < 1e-28). Participants with both CP and ACEs exhibited significantly reduced NAcc volume compared to those with neither condition (Cohen’s d = 0.39, p = 0.006). Other between-group differences were not statistically significant or exhibited negligible effect sizes (**Figure 3**b). Model assumptions of residual normality and homogeneity of variances were met in all analyses.

**Figure 3:**
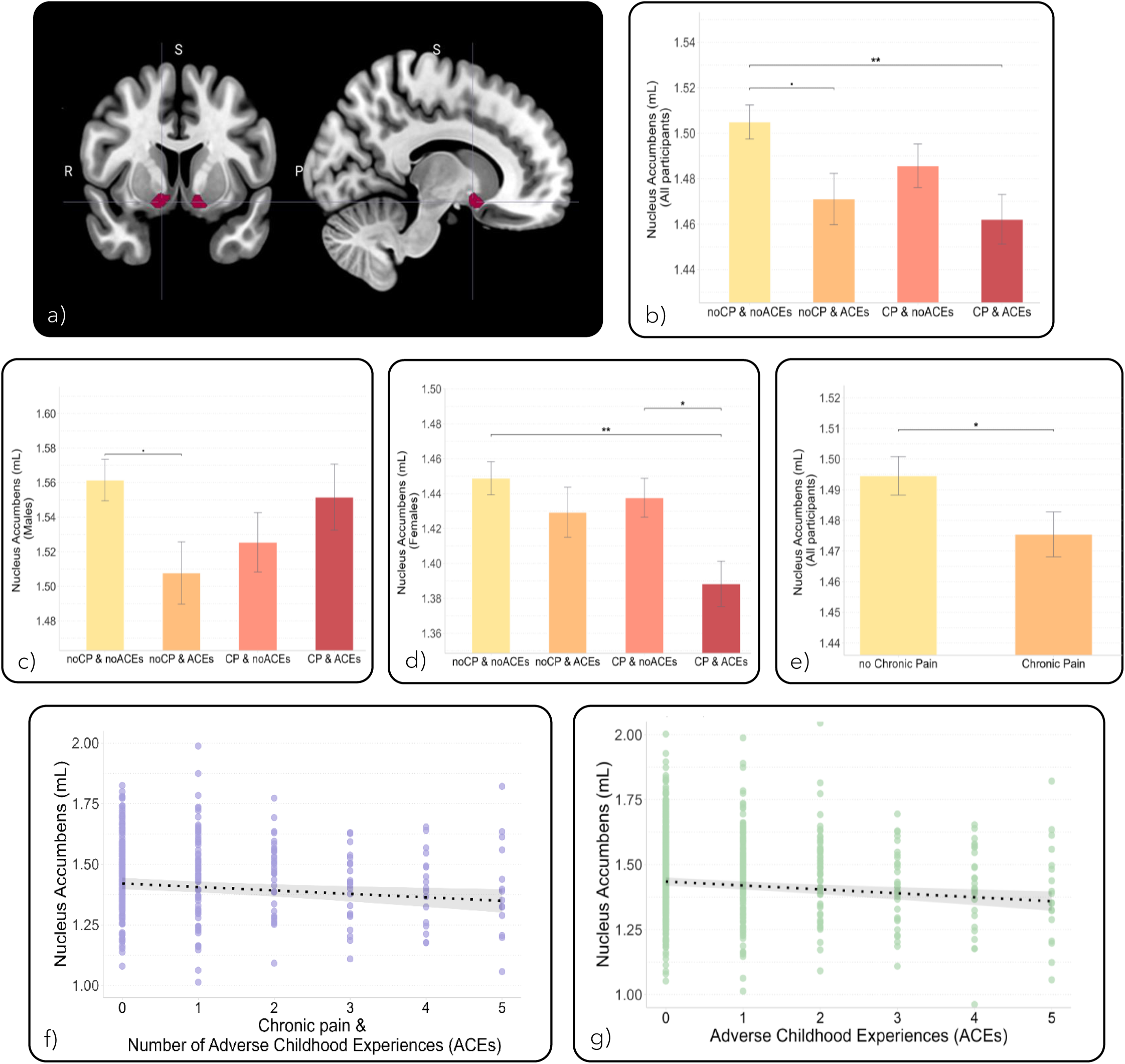
Associations between chronic pain, adverse childhood experiences (ACEs), and nucleus accumbens (NAcc) volume. a) Anatomical location of the nucleus accumbens (NAcc), the region of interest. b) NAcc volume by group: individuals without chronic pain or ACEs, those with ACEs only, those with chronic pain only, and those with both ACEs and chronic pain. ANCOVA model adjusted for age, sex, and site. c) Same as panel b but restricted to females. d) Same as panel b, but restricted to males. e) NAcc volume comparison between individuals with and without chronic pain. ANCOVA model adjusted for age, sex, and site. f) Linear regression showing the association between NAcc volume and ACEs exposure among individuals with chronic pain. g) Linear regression showing a negative association between NAcc volume and ACE exposure among all individuals. All models are adjusted for age, sex, and site.

Sex-stratified analyses revealed that these structural differences were more pronounced in females (**Figure 3**c and **Figure 3**d). Among women (N = 525), there was a significant main effect of CP/ACEs group (F(3,519) = 4.93, p = 0.002), with reduced NAcc volume observed in the CP & ACEs group compared to both the no CP/no ACEs group (Cohen’s d = 0.52, p = 0.001) and the CP-only group (Cohen’s d = 0.38, p = 0.021). In contrast, the effect among males (N = 322) was marginal (F(3,316) = 2.49, p = 0.06), with a trend-level difference between the no CP/no ACEs and no CP/ACEs groups (Cohen’s d = 0.37, p = 0.063). Age emerged as a robust covariate in all models, while imaging site showed no significant effect.

Independent ANCOVA models also indicated a significant main effect of CP status on NAcc volume (F(1,842) = 4.03, p = 0.045), with participants reporting CP having significantly lower volumes compared to those without CP (Cohen’s d = 0.24, p = 0.045) (**Figure 3**e). Linear regression analyses further showed that the combination of CP and the number of ACEs was predictive of reduced NAcc volume (β = –0.014, p = 0.006) (**Figure 3**f). Furthermore, ACEs, modelled alone as a continuous predictor, were associated with smaller NAcc volume (β = –0.015, p < 0.001) (**Figure 3**g). Both models accounted for key covariates and explained a similar proportion of variance (adjusted R² ≈ 0.22–0.23).

### Multivariate Prediction of Reward Signal

To determine whether clinical or structural variables can serve as predictors of functional reward activation, penalised regression models, including elastic net, LASSO, and ridge regression, were employed. (**Table 5**). Structural variation of grey matter volume showed a minimal and non-significant contribution to the prediction of functional reward signal (β = –0.012, p = .82 in standard regression; near-zero coefficients in LASSO and Elastic Net). Among all predictors, TIV and sex emerged as the most robust predictors, both negatively associated with reward activation and significant in the unpenalised model (TIV: β = –0.20, p = .003; sex: β = –0.17, p = .002). The best-performing model (elastic net) achieved modest performance (MSE = 0.96, R² = 0.041), retaining small coefficients for ACEs, QIDS, and CPG, which suggests subtle but consistent contributions from these clinical variables.

**Table 5:** Multivariate LASSO, Ridge and Elastic Net analyses examining the relationship between reward activation and the selected predictors. These models help handle multicollinearity and identify the most relevant variables while controlling for model complexity. Standard linear regression was also conducted for comparison. The table presents standardised beta coefficients, optimal λ, mean squared error (MSE) and R-squared values. Predictors include variables such as age, sex, scan site, total intracranial volume (TIV), the Quick Inventory of Depressive Symptomatology (QIDS), the Hospital Anxiety and Depression Scale (HADS), the chronic pain grade (CPG), the probability of grey matter in the basal ganglia and the adverse childhood experiences (ACEs) scores. The Elastic net model effectively predicts the reward activation signal in the basal ganglia, demonstrating significant negative associations for all the selected predictors. Probability Grey matter of the basal ganglia was not a significant predictor, suggesting that anatomical differences do not account for functional reward signal variation.

| Comparison of LASSO, Ridge, and Standard Linear Regression Coefficients<br>with MSE, R-squared, and $\lambda$ (Optimal) | | | | | |
| --- | --- | --- | --- | --- | --- |
|  | LASSO<br>Coefficient | Ridge<br>Coefficient | Elastic net<br>Coefficient | Standard linear<br>regression |  |
|  |  |  |  | Coefficient | p-value |
| (Intercept) | ~0 | ~0 | ~0 | ~0 | 1 |
| Structural variation<br>(basal ganglia) | -0.0144 | -0.0369 | -0.0144 | -0.012 | 0.821 |
| CPG | -0.0205 | -0.0251 | -0.0243 | -0.0278 | 0.548 |
| ACEs | -0.0247 | -0.0256 | -0.0295 | -0.0343 | 0.445 |
| QIDS | -0.0838 | -0.0588 | -0.0836 | -0.0847 | 0.173 |
| HADS | -0.0085 | -0.0254 | -0.0119 | -0.0141 | 0.809 |
| TIV | -0.1651 | -0.0901 | -0.1808 | -0.2013 | 0.003 ** |
| Sex | -0.1435 | -0.0833 | -0.1568 | -0.1736 | 0.002 ** |
| site | -0.0401 | -0.0244 | -0.0481 | -0.0573 | 0.215 |
| $\lambda$ (Optimal) | 0.0101 | 0.3555 | **0.0096** | NA | NA |
| MSE | 0.9581 | 0.9638 | **0.9574** | 0.957 | NA |
| R-squared | 0.0401 | 0.0345 | 0.0409 | 0.0412 | NA |

### Structural Equation Modelling: Linking Brain Structure, Reward Function, and Clinical Symptoms

The structural equation model (SEM) demonstrated good overall fit to the data, with a Comparative Fit Index (CFI) of 0.975 and Tucker-Lewis Index (TLI) of 0.930, both exceeding the recommended threshold of 0.90. The Root Mean Square Error of Approximation (RMSEA) was 0.063 (90% CI: 0.048–0.079), and the Standardised Root Mean Square Residual (SRMR) was 0.038, indicating acceptable error and residual variance (supplementary materials). The SEM accounted for 3.9% of the variance in reward-related activity, 3.1% in CP severity, and 1.6% in depressive symptoms, while variance explained for individual ACE subtypes ranged from 0.3% to 1.3%.

Structural variation in the basal ganglia positively predicted reward-related functional activity (β = 0.027, p = 0.005) (**Table 6**). In turn, reduced reward-related signal was associated with higher CP severity (β = –0.085, p = 0.039) and greater depressive symptoms (β = –0.121, p = 0.005). Reward-related activity also negatively predicted specific ACE subtypes, including sexual abuse (β = –0.090, p = 0.038) and physical neglect (β = –0.083, p = 0.052). Age was a consistent predictor across models, showing positive associations with multiple outcomes, including all ACE subtypes. Structural variation did not directly predict clinical outcomes or other types of ACEs at a statistically significant level, and no significant indirect (mediated) effects via the reward signal were detected (supplementary materials). Full path estimates, including all predictor and covariate relationships, are provided in the supplementary materials.

**Table 6:** Significant paths identified in the structural equation model linking structural brain variation, functional reward-related signal, and clinical outcomes. Estimates are unstandardised, and the final column reports standardised coefficients. Only significant paths (p < .05) or trends (p < .10) are reported here. Model covariates included age, sex, total intracranial volume (TIV), and site. Std. Est. refers to the standardised regression coefficient, allowing comparison across predictors on different scales. See Supplementary Table 9 for the full model. Significance: *** p < .001, ** p < .01, * p < .05, ^.^ p < .10

| Outcome | Predictor | Est. | SE | z | p | Std. Est. |
| --- | --- | --- | --- | --- | --- | --- |
| **Reward eigenvariate** | **Structural variation** | 0.414 | 0.810 | 0.51 | 0.005* | 0.027 |
|  | Sex | -0.255 | 0.091 | -2.81 | 0.005** | -0.173 |
|  | Age | 0.008 | 0.003 | 3.03 | 0.003** | 0.121 |
|  | TIV | -0.001 | 0.000 | -2.97 | 0.003** | -0.203 |
| **CPG** | **Reward eigenvariate** | -0.122 | 0.059 | -2.06 | 0.039* | -0.085 |
|  | Age | 0.009 | 0.004 | 2.37 | 0.018* | 0.102 |
|  | TIV | -0.001 | 0.000 | -2.37 | 0.018* | -0.131 |
| **QIDS** | **Reward eigenvariate** | -0.685 | 0.245 | -2.80 | 0.005** | -0.121 |
| **Physical Abuse** | Age | 0.009 | 0.004 | 2.37 | 0.018* | 0.040 |
| **Sexual Abuse** | **Reward eigenvariate** | -0.494 | 0.238 | -2.08 | 0.038* | -0.090 |
|  | Age | 0.009 | 0.004 | 2.37 | 0.018* | 0.027 |
| **Emotional Abuse** | Age | 0.009 | 0.004 | 2.37 | 0.018* | 0.028 |
| **Physical Neglect** | **Reward eigenvariate** | -0.274 | 0.141 | -1.94 | 0.052 . | -0.083 |
|  | Age | 0.009 | 0.004 | 2.37 | 0.018* | 0.045 |
| **Emotional Neglect** | Age | 0.009 | 0.004 | 2.37 | 0.018* | 0.024 |

## Discussion

In a large, community-based cohort comprising more than 800 well-phenotyped individuals, we demonstrate that ACEs, CP, and depression jointly impact the brain’s reward circuitry through both structural and functional mechanisms. Specifically, we identified reductions in NAcc GMV alongside blunted reward-related activation, changes that were dissociable and differentially predicted by clinical factors. Particularly, functional blunting of the reward signal, independently of structural changes, was associated with CP severity, depression symptoms, and exposure to sexual abuse, consistent with previous findings^12,24^ and in line with existing maltreatment literature^31,32^. Notably, CP grades, assessed 3 to 11 years prior to mood and ACEs measures, predicted long-term reductions in striatal reward activation, indicating a lasting impact on functional brain responses. These findings challenge assumptions that structural loss underlies functional impairments and demonstrate distinct, interacting neurobiological pathways.

A reduction in NAcc GMV was observed in participants with CP compared to those without, primarily driven by effects in females, consistent with previous literature^33^. Crucially, the effect was present only when CP co-occurred with ACEs; neither exposure alone produced significant change. These findings suggest that NAcc structural alterations may emerge more robustly under combined exposure, particularly in women, indicating sex-specific vulnerability to early adversity and pain-related neuroplasticity.

The blunted reward response observed may reflect dopaminergic dysfunction, a mechanism supported by clinical and preclinical models. While the NAcc is well established in reward processing, mood regulation, and addiction, its role in pain modulation is increasingly recognised^33^. Reduced NAcc activation during pain onset^30,34^ and abnormalities in reward-related processing across chronic pain conditions^24,35^ support its dual role in nociception and motivation. CP has been associated with impaired dopaminergic neurotransmission^36–38^, and affective alterations^39^ characteristic of anhedonia and depression^40^. Consistent with this, ventral striatal hypoactivation, including in the NAcc, is well documented in major depressive disorder^12,22^, and this is consistent with our results.

ACEs may contribute to similar alterations through stress-induced dopaminergic dysregulation. Chronic stress reduces dopaminergic neurons in the VTA^41^ and alters activity in NAcc and medial prefrontal circuits^41,42^. Rodent studies show that early-life adversity disrupts normal dopaminergic development⁴⁵, alters reward-related behaviour^43,44^, and enhances opioid-seeking following early neglect^45^. These models align with our finding that sexual abuse-related ACEs predict reduced reward activation and suggest that both early adversity and CP may act on similar neural substrates via stress and dopaminergic mechanisms. Future work should examine these pathways longitudinally in humans to identify shared and dissociable biological signatures of risk.

Despite these structural and clinical associations, regularised regression models (Elastic Net, LASSO, ridge) showed that GMV contributed minimally to predicting reward-related activation. Clinical variables, including ACEs, QIDS scores, and CPG, showed small but stable contributions across models. This suggests that functional reward alterations are not directly driven by basal ganglia volume, indicating a partial decoupling between structure and function. Although structural reductions are seen in individuals with CP and ACEs^33^, they do not directly correlate with variations in functional reward signals^24,31,32^. This highlights partially distinct pathways through which early adversity, pain, and mood may influence reward processing, with sex and brain size playing a larger role in functional variability than structural changes.

Structural equation modelling further supported this dissociation, while basal ganglia volume weakly predicted activation (β = 0.027, p = 0.005), only functional reward signals were significantly associated with depressive symptoms, CP severity, and sexual abuse-related ACEs, independent of structural variation. These findings suggest that adversity and pain have overlapping yet distinct pathways that impact the brain, with functional signals being more closely linked to current affective and motivational states.

There are some limitations to this study: Retrospective assessment of ACEs introduces potential recall bias, with evidence suggesting modest congruence at best between retrospective and prospective measures, and CP and depression may themselves impact the retrospective recall of ACEs^46^. CP was measured before imaging and mood assessments; while temporally misaligned, this allows inference about the lasting effects of earlier pain exposure. The reward task was brief to accommodate other data collection, but it allowed for comparison against a neutral baseline rather than punishment, thereby aiding in interpretability. Finally, the sample was non-clinical and community-based, which strengthens generalisability but may underestimate effects present in clinical populations.

Collectively, these findings enhance our understanding of how adversity and pain influence the human reward system. Rather than reflecting a single continuum of dysregulation, reward-related brain changes appear to arise via distinct but converging mechanisms. Structural alterations, particularly in the context of ACEs and CP, may reflect cumulative or developmental effects on brain morphology. In contrast, functional blunting of reward signals appears more sensitive to current affective symptoms and motivational dysregulation. Our results support a dual-mechanism model and underscore the need for mechanistically informed interventions that target both structural vulnerability and dynamic functional changes. Understanding how adversity and pain interact to affect brain circuitry is critical for developing early interventions that could improve outcomes for individuals with these comorbidities, particularly in clinical settings where such conditions are prevalent.

## Methods

### Study Design

This was a secondary data analysis of a pre-existing cohort: The Generation Scotland Scottish Family Health Study (GS:SFHS) is a longitudinal cohort involving approximately 24,000 community-based participants^47^, initially recruited between 2006 and 2011^47^. Participants in the Stratifying Resilience and Depression Longitudinally (STRADL) study, were recruited from the GS:SFHS 3 to 11 years later^48,49^. Consent to link participant data to NHS records was obtained during the original GS:SFHS (05/S1401/89). All later procedures for the STRADL study were conducted under a separate but related ethics application (14/SS/0039). This study uses the STRADL dataset, approved by the Generation Scotland Access Committee (GSAC) under application GS21383. The Chronic Pain Advisory Group (CPAG) is a patient advisory panel, independent of the GS:SFHS cohort, that shares insights based on lived experiences. CPAG participated throughout the research cycle.

### Participants and Procedures

The GS:SFHS study includes socio-demographic and clinical information collected at study entry (for more information see supplementary materials). CP was assessed at the time of entry to the GS:SFHS study (2006-2011) using a validated Chronic Pain Definition (CDQ) questionnaire^50^ and the 7-item Chronic Pain Grade (CPG) questionnaire^51^. The CPG questionnaire was used because it provides a comprehensive assessment of CP, considering both pain intensity and its interference with daily activity. We analysed a linked subset of the GS:SFHS participants who completed the CDQ questionnaires and who also took part in the STRADL study^48,49^.

STRADL participants completed face-to-face clinical and neuropsychological assessments, including basic demographics, health and lifestyle, physical measures, psychiatric questionnaires, cognitive tests, laboratory sample collection and structural and functional brain magnetic resonance imaging (MRI) scanning, at the Universities of Aberdeen and Dundee. A research version of the Structured Clinical Interview for Diagnostic and Statistical Manual of Mental Disorders (DSM-IV) (SCID)^52^ was utilised to assess prior indications of mood disorders and any occurrences of manic or hypomanic episodes, using the DSM-IV-TR^53^ for the diagnostic criteria. The Quick Inventory of Depressive Symptomatology (QIDS)^54^ was administered to assess depression severity. A restricted set of ACEs, defined as childhood or adolescent abuse or neglect, was assessed using the Childhood Trauma Questionnaire short form (CTQ-SF)^55^ in the STRADL study. This is a 28-item quantitative and retrospective inventory with substantial or excellent internal consistencies for most scales in both community and clinical samples that assesses histories of abuse and neglect, with subscales for emotional, physical, and sexual abuse and emotional and physical neglect.

T1-weighted structural brain images and functional MRI data were acquired as part of STRADL at the Universities of Dundee and Aberdeen. In Aberdeen, participants were imaged on a 3T Philips Achieva TX-series MRI system (Philips Page 10 of 31 Healthcare, Best, Netherlands) with a 32-channel phased-array head coil and a back facing mirror (software version 5.1.7; gradients with maximum amplitude 80 mT/m and maxi-mum slew rate 100 T/m/s). For the functional MRI acquisition in Aberdeen a projector and “Presentation” (Neurobehavioural Systems Inc, Berkeley, CA, USA) version 18.1 were used with a repetition time 1.56 s, echo time of 26 ms, flip angle 70◦, field of view 217 mm, matrix 64×64, 32 axial slices were used. In Dundee, participants were scanned using a Siemens 3T Prisma-FIT (Siemens, Erlangen, Germany) with 20 channel head and neck phased array coil and a back facing mirror (Syngo E11, gradient with max amplitude 80 mT/m and maximum slew rate 200 T/m/s). A magnetic resonance compatible LCD screen was used to display fMRI (NordicNeuroLab, Bergen, Norway) task stimuli using “Presentation” version 20.0 with a repetition time of 1.56 s, echo time 22 ms, flip angle 70◦, field of view 217 mm, matrix 64×64 and 32 axial slices.

An established probabilistic reward learning fMRI task^56^ with 66 trials was completed by STRADL participants^12^. On each trial, one of two stimuli (blue or yellow squares) was chosen, and each stimulus was associated with different reward probabilities: 80% for a yellow square and 20% for a blue square. Feedback was provided on participants’ choices by showing points on the computer screen: 100 points for a ‘win’ i.e., reward and 0 points for ‘no win’ i.e., no reward. Each trial was started with a cue indicating whether the computer would choose a square to select, in which case participants had to follow the choice made by the computer, or whether participants would be allowed to make their own choice of square. The number of trials was split into 33 ‘choice’ and 33 ‘no-choice’ randomly presented trials, where the timing of presentation was varied (jittered) to allow for identification of the signals of interest. Participants were included in the analysis if they had completed the CTQ-SF, QIDS and CDQ questionnaires, and had good quality fMRI images available.

### Task-based fMRI analyses

Neuroimaging analyses were conducted using SPM12 (Statistical Parametric Mapping)^57^. An event-related design was used for the first-level analysis. A first-level general linear model (GLM) design matrix included two columns of possible outcomes: reward during a choice or no-choice trial; and no-reward during a choice or no-choice trial. As covariates of no interest, the six rigid body motion realignment parameter estimates during pre-processing were included. Events were modelled as truncated delta functions and convolved with the SPM12 canonical haemodynamic response function without time or dispersion derivatives. Participants who still had a signal dropout from the predefined areas of interest, such as the basal ganglia, were excluded from the second-level analysis.

Contrast estimates (e.g., activation of reward signal during choice and no choice conditions compared to no reward during choice and no choice conditions) from each subject were taken to the second-level analysis, which treated inter-subject variability as a random effect to account for inter-individual variance. This allowed tests for correlations between fMRI data and depression scores (QIDS), ACEs sub-scores (CTQ) and CPG. A cluster extent threshold which involved Monte Carlo simulations was used^58,59^. Multiple comparisons of effects linked to depressive symptom severity, chronic pain severity and ACEs were controlled using a whole brain cluster corrected threshold of P*<*0.001, comprising a simultaneous requirement for a P*<*0.05 voxel threshold and *>*130 contiguous supra-threshold voxels. Based on previous literature, we selected the basal ganglia mask for the ROI analysis^11^. Volumes of interest masks for the basal ganglia were created using the WFU PickAtlas toolbox^60^. The REX toolbox^61^ was used to extract the first eigenvariate values of the beta values of the contrast estimates, the de-activation of reward compared to no-reward during choice and no choice conditions, from each subject.

### Structural MRI analyses

Image pre-processing for the structural T1-weighted images was performed using the Computational Anatomy Toolbox (CAT12)^62^, starting with the segmentation process. Tissue Probability Maps (TPMs) are utilised to initialise the segmentation process. The TPMs serve as initial estimates to guide the segmentation algorithm during the tissue classification process. For spatial registration, Geodesic Shooting^63^ was performed for its advanced approach of calculating geodesic distances to transform individual brain images into a common space. It generates transformations that ensure anatomical consistency between brain images while minimising geometric distortions. The sample homogeneity was assessed after the segmentation process by utilising the weighted overall image quality and the quartic mean Z-score. A low ratio indicates a good quality after the pre-processing, meaning a high rated weighted overall image quality and high quartic mean Z-score. Additionally, the total intracranial volume (TIV) was calculated to be used later as a covariate. This led to the final pre-processing step, smoothing the functional images using an 8mm full-width at half-maximum Gaussian kernel.

To compare tissue volumes between different groups based on the voxels, a statistical analysis, a VBM analysis, was performed for the already pre-processed and smoothed images. This was done by utilizing the general linear model (GLM) method to identify the regions of significant differences by generating statistical maps that indicate regions where significant differences exist. The grey matter images from each participant treated inter-subject variability as a random effect to account for inter-individual variance, such as age. Global normalisation was also employed in order to account for different brain sizes by calculating the total intracranial volume and using it for proportional scaling. After estimating the GLM, comparison between different groups or correlation of the brain structure with the clinical variables was performed. More specifically, the correlation between sub-scores of CTQ, chronic pain severity, the comorbidity of the two with the grey matter was performed.

Due to the large number of comparisons conducted in voxel-based morphometry (VBM) analysis, multiple comparison corrections were applied to control the potential risk of false positive results. This was performed by utilising a cluster-extended threshold based on Monte Carlo simulations, with the threshold of significance specified at p< 0.05 and corrected at a whole-brain cluster level^58,59,64^. The calculation was carried out as previously described^64^, using 10,000 iterations. A cluster size threshold of 144 voxels was determined, ensuring that the probability of encountering clusters of this size or larger was less than p < .05 across the entire brain^64^.

A Region of Interest (ROI) analysis was performed in predefined brain regions based on their anatomical and functional relevance, thereby reducing multiple comparison issues associated with whole-brain analysis. More specifically, these regions were selected based on results from our previous systematic review and meta-analysis^11^. The volumes of specific regions of interest were extracted in CAT12. After that, in R-studio^65^a comparison of the volumes among the various groups was performed (using the same groups as those in the Voxel-Based Morphometry (VBM) analysis), corrected for multiple comparisons using the False Discovery Rate (FDR) with a threshold of significance set at p < 0.05. Additionally, a one-way analysis of covariance (ANCOVA) was conducted to examine group differences in NAcc volume across four groups defined by the presence or absence of CP and ACEs. The primary independent variable was group (CP/ACEs status), and the dependent variable was regional brain volume. To control for potential confounding effects, age, sex and imaging site were included as covariates in the model. Prior to interpreting ANCOVA results, model assumptions were assessed. The normality of residuals was evaluated using the Shapiro–Wilk test, and homogeneity of variance was tested using Levene’s test. For significant main effects, post hoc pairwise comparisons were performed using Tukey-adjusted p-values. Standardized effect sizes (Cohen’s d) were calculated for each group comparison, and interpreted using conventional thresholds (small, medium, large). Generalized eta squared (GES) was computed for all main effects to quantify the proportion of variance explained by each factor.

### Regularized Regression Analysis

We implemented a comprehensive regression analysis pipeline to predict the outcome variable reward-related basal ganglia functional signal from a set of predictors including structural variation of the basal ganglia, clinical measures (e.g., chronic pain severity, depression symptom severity QIDS, anxiety symptom severity HADS, ACEs), and demographic covariates (TIV — total intracranial volume, Sex, and site). The structural variation in the basal ganglia was of primary interest and was summarised by extracting the first eigenvariate from a bilateral ROI using the Rex toolbox in SPM12, based on voxel-based morphometry (VBM) data. This approach allowed us to test whether inter-individual differences in functional activation were attributable to corresponding structural variation. Continuous predictor variables and outcomes were standardised (mean-centred and scaled to unit variance) before fitting the regularised regression models to ensure comparability of effect sizes and appropriate penalisation. Categorical covariates, such as sex and site, were encoded numerically or as factors, as appropriate for each analysis.

Four models were fitted and compared:

- LASSO regression (L1 penalty, alpha = 1) performs variable selection by shrinking some coefficients exactly to zero.
- Ridge regression (L2 penalty, alpha = 0) shrinks coefficients towards zero but does not set them exactly to zero.
- Elastic Net regression (mixed L1 and L2 penalties, alpha = 0.5) balances variable selection and coefficient shrinkage.
- Standard linear regression without penalisation for baseline comparison.

These regularised regressions are particularly important as they effectively handle multicollinearity, which is crucial given the likely covariance among predictors such as pain, ACEs, age, and QIDS/HADS. Model fitting was performed using the glmnet package in R, employing 10-fold cross-validation to select the optimal regularisation parameter (lambda), thereby minimising the mean squared error (MSE). The models were fit on standardised predictors and outcome variables. Model performance was evaluated by computing mean squared error (MSE) and coefficient of determination (R²) on the training data. The best-performing regularised model (lowest MSE) was identified, and coefficients were refitted on the original (unstandardised) data using the optimal lambda to enable interpretation on the raw scale.

### Structural Equation Modelling (SEM) Analysis

To investigate potential mediating pathways linking structural variation in the basal ganglia to chronic pain severity, psychological symptoms, and ACE subdomains via reward-related functional activation, we employed structural equation modelling (SEM) using the lavaan package in R. The model adjusted for relevant covariates: age, sex, total intracranial volume (TIV), and imaging site. Paths from both the structural and functional variables to each outcome were estimated, along with indirect effects via the functional mediator (i.e., mediation pathways) and total effects (sum of direct and indirect effects). Variables were scaled and centred before model estimation to aid interpretability. Site was entered as a categorical variable. Model estimation used maximum likelihood with bootstrapped standard errors (1,000 resamples), and model fit was evaluated using established indices, including the Comparative Fit Index (CFI), Tucker-Lewis Index (TLI), and Root Mean Square Error of Approximation (RMSEA). R² values were computed for each endogenous variable to assess the proportion of explained variance.

## Data Sharing

Data can be accessed upon reasonable request. Researchers can apply for access to Generation Scotland data via the website (https://www.ed.ac.uk/generation-scotland/for-researchers).

## Acknowledgements

The study is part of the E-PaiD PhD project, funded by the TENOVUS Scotland Research Studentship, ref T20-18. LC, BS, DS, TH and GA are members of the Advanced Pain Discovery Platform. LC, DS, TH and GA are also members of the Consortium Against Pain Inequality which received funding from UKRI and Versus Arthritis Grant: MR/W002566/1. Generation Scotland received core support from the Chief Scientist Office of the Scottish Government Health Directorates [CZD/16/6] and the Scottish Funding Council [HR03006] and is currently supported by the Wellcome Trust [216767/Z/19/Z]. Imaging substudy was funded through a Wellcome Trust (Wellcome Trust Strategic Award “STratifying Resilience and Depression Longitudinally” (STRADL)) [104036/Z/14/Z].

